# The Role of Social Prescribing in Alleviating Social Isolation and Loneliness in Older Adults: A Systematic Review Protocol

**DOI:** 10.1101/2025.07.07.25331068

**Authors:** Ravi Shankar, Fiona Devi, Xu Qian

## Abstract

**Background:** Social isolation and loneliness are significant public health concerns, particularly among older adults. Social prescribing, a non-medical intervention that links individuals to community-based support and activities, has gained attention as a potential solution. However, the effectiveness of social prescribing in addressing social isolation and loneliness in older adults remains unclear.

**Objective:** This systematic review aims to examine the current evidence on the role of social prescribing in alleviating social isolation and loneliness among older adults.

**Methods:** A comprehensive search will be conducted in multiple electronic databases, including PubMed, Web of Science, Embase, CINAHL, MEDLINE, The Cochrane Library, PsycINFO, and Scopus, from inception to July 2025. Gray literature sources and reference lists of included studies will also be searched. The PICO framework will be used to define the population, intervention, comparator, and outcomes of interest. Eligible studies will include randomized controlled trials, quasi-experimental studies, and observational studies that assess the impact of social prescribing on social isolation and loneliness in adults aged 60 years and above. Two reviewers will independently screen titles, abstracts, and full-text articles using Covidence software. Data extraction and risk of bias assessment will be conducted using standardized tools. A narrative synthesis will be conducted to summarize the findings, with subgroup analyses based on study design, intervention characteristics, and participant demographics.

**Discussion:** This systematic review will provide a comprehensive evaluation of the existing evidence on the effectiveness of social prescribing in addressing social isolation and loneliness among older adults. The findings will inform healthcare providers, policymakers, and researchers about the potential of social prescribing as a non-medical intervention and guide future research and implementation efforts.

**PROSPERO Registration:** CRD42024614147

## Introduction

## Background

Social isolation and loneliness are increasingly recognized as significant public health concerns, particularly among older adults. Social isolation refers to the objective lack of social contacts and interactions, while loneliness is a subjective feeling of being alone or isolated [1]. Both social isolation and loneliness have been associated with adverse physical and mental health outcomes, including increased risk of cardiovascular disease, cognitive decline, depression, and mortality [2,3].

The prevalence of social isolation and loneliness among older adults is substantial. A systematic review by Ong et al. [4] found that the prevalence of social isolation ranged from 7% to 24% among adults aged 60 years and above, while the prevalence of loneliness ranged from 5% to 43%. With the global population aging rapidly, addressing social isolation and loneliness in older adults has become a pressing issue.

Several factors contribute to the increased risk of social isolation and loneliness among older adults. As individuals age, they may experience life transitions such as retirement, loss of spouse or friends, and reduced mobility, which can lead to a decline in social connections and support [5]. Additionally, age-related health conditions and disabilities can limit older adults’ ability to participate in social activities and maintain relationships [6].

The COVID-19 pandemic has further exacerbated the issue of social isolation and loneliness among older adults. Physical distancing measures and restrictions on social gatherings have disproportionately affected older adults, who are at higher risk of severe illness from the virus [7]. The pandemic has highlighted the need for innovative interventions to address social isolation and loneliness, particularly in the context of public health emergencies.

### Social Prescribing

Social prescribing is a non-medical intervention that links individuals to community-based support and activities to improve their health and well-being [8]. It involves healthcare professionals, typically primary care providers, referring patients to a link worker or community navigator who assesses their needs and connects them to appropriate services, such as social activities, exercise groups, art classes, and volunteering opportunities [9].

The concept of social prescribing is based on the recognition that social determinants of health, such as social support, social engagement, and access to community resources, play a crucial role in an individual’s overall health and well-being [10]. By addressing these social determinants, social prescribing aims to complement traditional medical interventions and provide a holistic approach to healthcare.

Social prescribing has gained momentum in recent years as a potential solution to address the social determinants of health and reduce the burden on healthcare systems. In the United Kingdom, social prescribing has been endorsed by the National Health Service (NHS) as part of its Long Term Plan [11]. The NHS has committed to expanding social prescribing services, with a goal of referring 900,000 people to social prescribing by 2023/24 [12].

Other countries have also shown growing interest in social prescribing. In Canada, the Alliance for Healthier Communities has launched the Social Prescribing in Ontario project, which aims to build capacity for social prescribing in primary care settings [13]. In Australia, the Royal Australian College of General Practitioners has recognized social prescribing as a promising approach to address the social determinants of health and has called for its integration into primary care [14].

### Rationale for the Review

Despite the increasing attention on social prescribing, its effectiveness in alleviating social isolation and loneliness among older adults remains unclear. Previous reviews have explored the impact of social prescribing on various health outcomes, but they have not specifically focused on social isolation and loneliness in older adults [15,16].

A systematic review by Bickerdike et al. [15] examined the evidence on the effectiveness of social prescribing for a range of health and well-being outcomes. While the review included studies that measured social isolation and loneliness, it did not provide a comprehensive analysis of the impact of social prescribing on these specific outcomes in older adults.

Similarly, a systematic review by Chatterjee et al. [16] investigated the effectiveness of non-clinical community interventions, including social prescribing, on health and well-being outcomes. Although the review included studies with older adult populations, it did not specifically examine the impact of social prescribing on social isolation and loneliness.

Given the significant burden of social isolation and loneliness among older adults and the potential of social prescribing to address these issues, a comprehensive review of the evidence is warranted. This systematic review aims to fill this gap by focusing specifically on the role of social prescribing in alleviating social isolation and loneliness in older adults.

### Objectives

The primary objective of this systematic review is to examine the current evidence on the role of social prescribing in alleviating social isolation and loneliness among older adults. Specifically, the review aims to:

1. Assess the effectiveness of social prescribing interventions in reducing social isolation and loneliness in adults aged 60 years and above.
2. Identify the characteristics of effective social prescribing interventions, such as the type of activities, duration, and intensity.
3. Explore the potential moderators of intervention effectiveness, such as participant demographics, health status, and social support.
4. Evaluate the quality of the existing evidence and identify gaps in the literature to guide future research.

## Methods

### Protocol and Registration

This systematic review protocol has been registered with the International Prospective Register of Systematic Reviews (PROSPERO) to ensure transparency and reduce the risk of duplication (PROSPERO registration number: CRD42024614147). The protocol will be reported in accordance with the Preferred Reporting Items for Systematic Review and Meta-Analysis Protocols (PRISMA-P) guidelines [17].

### Eligibility Criteria

The PICO (Population, Intervention, Comparator, Outcomes) framework will be used to define the eligibility criteria for this systematic review.

### Population

Adults aged 60 years and above, regardless of their health status, living in community settings. Studies focusing on specific subgroups, such as those with chronic conditions or dementia, will be included if the mean age of participants is 60 years or above.

### Intervention

Social prescribing interventions that involve healthcare professionals referring individuals to community-based support and activities. The interventions may vary in terms of the type of activities, duration, and intensity. Examples of social prescribing activities include:

- Social activities: group meetings, clubs, social events
- Physical activities: exercise classes, walking groups, sports teams
- Arts and cultural activities: art classes, music groups, museum visits
- Nature-based activities: gardening, horticultural therapy, outdoor activities
- Learning and skill development: language classes, computer training, cooking classes
- Volunteering and community involvement: volunteering opportunities, community projects

### Comparator

No intervention, usual care, or alternative interventions that do not involve social prescribing, such as:

- Waitlist control
- Standard medical care
- Pharmacological interventions
- Psychological therapies
- Other non-pharmacological interventions (e.g., cognitive training, physical exercise)

### Outcomes

The primary outcomes of interest are social isolation and loneliness, measured using validated scales or instruments, such as:

- UCLA Loneliness Scale [18]
- De Jong Gierveld Loneliness Scale [19]
- Lubben Social Network Scale [20]
- Duke Social Support Index [21]
- Campaign to End Loneliness Measurement Tool [22]

Secondary outcomes may include a range of psychosocial and health-related measures such as health-related quality of life, depression, anxiety, social support, social participation, social connectedness, wellbeing, self-esteem, healthcare utilization, and adverse events.

### Study Design

Randomized controlled trials (RCTs), quasi-experimental studies, and observational studies (cohort, case-control, and cross-sectional) will be included. Qualitative studies, case reports, and opinion pieces will be excluded.

### Language

Studies published in English will be included. If relevant studies in other languages are identified, attempts will be made to translate them.

### Information Sources

A comprehensive search will be conducted across several electronic databases from their inception to July 2025, including PubMed, Web of Science, Embase, CINAHL, MEDLINE, the Cochrane Library, PsycINFO, and Scopus. To capture relevant gray literature, additional searches will be carried out in ProQuest Dissertations & Theses Global, Open Grey, and Google Scholar, as well as on the websites of relevant organizations such as NHS England, the Social Prescribing Network, and the National Academy for Social Prescribing. Furthermore, the reference lists of all included studies and relevant reviews will be hand-searched to identify any additional eligible studies.

### Search Strategy

The search strategy will be developed in collaboration with a medical librarian and will include a combination of keywords and controlled vocabulary terms (e.g., Medical Subject Headings [MeSH]) related to social prescribing, social isolation, loneliness, and older adults. The search terms will be adapted for each database. An example search string for PubMed is provided below:

(“social prescribing” OR “community referral” OR “link worker” OR “community navigator”) AND (“social isolation” OR loneliness OR “social support” OR “social network” OR “social participation” OR “social connectedness”) AND (“older adults” OR elderly OR aged OR “older people” OR “aging population”)

The search string will be further refined to include relevant study design filters, such as the Cochrane Highly Sensitive Search Strategy for identifying randomized trials [23].

The search results will be exported to Covidence, a web-based software platform for systematic review management, for duplicate removal and screening.

### Study Selection

Two reviewers will independently screen the titles and abstracts of the retrieved studies against the eligibility criteria using Covidence. Full-text articles of potentially eligible studies will be obtained and independently assessed by the two reviewers. Any disagreements will be resolved through discussion or by consulting a third reviewer. The reasons for exclusion will be recorded at the full-text stage. The study selection process will be documented using a PRISMA flow diagram [24].

### Data Extraction

A standardized data extraction form will be developed and piloted on a sample of included studies. Two reviewers will independently extract data from the included studies using Covidence. The extracted data will include:

- Study characteristics (authors, year, country, study design, sample size, study duration)
- Participant characteristics (age, gender, ethnicity, health status, living arrangements, socioeconomic status)
- Intervention details (type of activities, frequency, duration, intensity, mode of delivery, link worker involvement, theoretical framework)
- Comparator details (type of control, description)
- Outcome measures (primary and secondary outcomes, measurement tools, time points)
- Results (effect sizes, confidence intervals, p-values, findings from subgroup and sensitivity analyses)
- Funding sources and conflicts of interest

Any discrepancies in the extracted data will be resolved through discussion or by consulting a third reviewer.

### Risk of Bias Assessment

The risk of bias in the included studies will be assessed using appropriate tools based on the study design. For RCTs, the Cochrane Risk of Bias tool (RoB 2) will be used [25]. The RoB 2 tool assesses bias in five domains: randomization process, deviations from intended interventions, missing outcome data, measurement of the outcome, and selection of the reported result.

For non-randomized studies, the Risk Of Bias In Non-randomized Studies of Interventions (ROBINS-I) tool will be employed [26]. The ROBINS-I tool assesses bias in seven domains: confounding, selection of participants, classification of interventions, deviations from intended interventions, missing data, measurement of outcomes, and selection of the reported result.

Two reviewers will independently assess the risk of bias, and any disagreements will be resolved through discussion or by consulting a third reviewer. The results of the risk of bias assessment will be presented in a summary table and considered in the interpretation of the findings.

### Data Synthesis

A narrative synthesis will be conducted to summarize the findings of the included studies. The synthesis will be structured around the primary and secondary outcomes, with subgroup analyses based on study design, intervention characteristics, and participant demographics.

If sufficient data are available and the studies are homogeneous in terms of design and outcomes, a meta-analysis will be considered. The choice between a fixed-effect or random-effects model will be based on the assessed heterogeneity among studies. Statistical heterogeneity will be assessed using the I^2^ statistic, with values above 50% indicating substantial heterogeneity [27].

If a meta-analysis is not feasible due to heterogeneity or insufficient data, the findings will be synthesized narratively. The narrative synthesis will involve organizing the studies into logical categories, examining patterns and relationships within and between studies, and identifying factors that may explain variations in the findings.

Subgroup analyses will be conducted, if possible, to explore the potential moderators of intervention effectiveness. Subgroups may include:

- Age groups (e.g., 60-69, 70-79, 80+)
- Gender
- Living arrangements (e.g., living alone, living with others)
- Health status (e.g., chronic conditions, frailty)
- Intervention characteristics (e.g., type of activity, duration, intensity)
- Study design (e.g., RCTs, quasi-experimental studies, observational studies)

Sensitivity analyses will be performed to assess the robustness of the findings by excluding studies with high risk of bias or other potential sources of heterogeneity.

Publication bias will be assessed using funnel plots and Egger’s test [28], if there are sufficient studies (at least 10) for a specific outcome.

The certainty of the evidence for each outcome will be assessed using the Grading of Recommendations, Assessment, Development, and Evaluation (GRADE) approach [29]. The GRADE assessment will consider factors such as the risk of bias, inconsistency, indirectness, imprecision, and publication bias. The certainty of evidence will be rated as high, moderate, low, or very low.

## Discussion

This systematic review protocol outlines the methods for a comprehensive evaluation of the current evidence on the role of social prescribing in alleviating social isolation and loneliness among older adults. The review will address a significant public health issue and inform policy and practice decisions regarding the use of social prescribing as a non-medical intervention.

The strengths of this review include the systematic search strategy, the use of the PICO framework to define eligibility criteria, the inclusion of a broad range of study designs, and the assessment of the risk of bias and certainty of the evidence using standardized tools. Additionally, the use of Covidence software for study selection and data extraction will enhance the efficiency and transparency of the review process.

However, potential limitations should also be acknowledged. The heterogeneity in the interventions and outcome measures may limit the ability to conduct a meta-analysis and draw firm conclusions. The inclusion of non-randomized studies may introduce bias and confounding factors that could affect the validity of the findings. Moreover, the inclusion of only English language studies may introduce language bias and limit the generalizability of the findings to non-English speaking populations.

Despite these limitations, this systematic review will provide a valuable synthesis of the existing evidence and identify gaps in the literature to guide future research. The findings will be relevant to healthcare providers, policymakers, and researchers interested in addressing social isolation and loneliness among older adults. The review will also contribute to the growing body of evidence on the potential of social prescribing as a non-medical intervention to improve health and well-being.

The dissemination of the review findings will involve multiple strategies to reach a wide audience. The review will be published in a peer-reviewed journal and presented at relevant conferences and workshops. A lay summary of the findings will be prepared for dissemination to the general public, older adults’ organizations, and community groups. The review team will also engage with policymakers and healthcare organizations to inform decision-making and promote the integration of social prescribing into healthcare systems.

In conclusion, this systematic review protocol presents a rigorous and transparent methodology to examine the effectiveness of social prescribing in alleviating social isolation and loneliness among older adults. The findings will have important implications for policy and practice and advance our understanding of the role of social prescribing in promoting healthy aging. By addressing a significant public health issue and providing evidence-based recommendations, this review has the potential to improve the health and well-being of older adults and contribute to the development of person-centered, community-based interventions.

## Data Availability

All data produced in the present study are available upon reasonable request to the corresponding author.

## APPENDIX A Data Extraction Form

**Table 1.**

Appendix A: Data Extraction Form
| Category | Items |
| --- | --- |
| **Study Characteristics** | First author<br>Year of publication<br>Country<br>Study design (RCT, quasi-experimental, observational)<br>Sample size<br>Study duration |
| **Participant Characteristics** | Age (mean, range)<br>Gender (% female)<br>Ethnicity<br>Health status<br>Living arrangements<br>Socioeconomic status |
| **Intervention Details** | Type of social prescribing activities<br>Frequency of activities<br>Duration of intervention<br>Intensity of intervention<br>Mode of delivery (face-to-face, remote, mixed)<br>Link worker involvement (yes/no, details)<br>Theoretical framework (if applicable) |
| **Comparator Details** | Type of control (no intervention, usual care, alternative intervention)<br>Description of control condition |
| **Outcomes** | **Primary outcomes** (e.g., social isolation, loneliness)<br>– Measurement tools used<br>– Time points assessed<br>**Secondary outcomes** (e.g., health-related quality of life, depression, anxiety, social support, social participation, social connectedness, well-being, self-esteem, healthcare utilization, adverse events)<br>– Measurement tools used<br>– Time points assessed |
| **Results** | Effect sizes<br>Confidence intervals<br>p-values<br>Findings from subgroup analyses<br>Findings from sensitivity analyses |
| **Other Information** | Funding sources<br>Conflicts of interest<br>Additional notes |

